# Serum autotaxin is associated with DHEAS and predicts longitudinal cognitive changes in older women: Analysis of the Arao cohort study

**DOI:** 10.64898/2026.03.17.26348449

**Authors:** Shu Sun, Naoto Kajitani, Kazuhiro Yoshiura, Manabu Makinodan, Minoru Takebayashi

**Affiliations:** Department of Psychiatry and Neuroscience, Center for Metabolic Regulation of Healthy Aging, Faculty of Life Sciences, Kumamoto University, Kumamoto, Japan; Department of Neuropsychiatry, Kumamoto University, Kumamoto, Japan; Department of Frailty Research, Center for Gerontology and Social Science, Research Institute, National Center for Geriatrics and Gerontology, Obu, Japan

**Keywords:** Autotaxin, DHEAS, LPA, Community Dwelling, Cognition

## Abstract

**Aim:** To examine the determinants of serum autotaxin (ATX) levels in community-dwelling older adults, focusing specifically on endogenous adrenal steroids, and to investigate the cross-sectional and longitudinal associations between serum ATX levels, cognitive function, and depressive symptoms.

**Methods:** Data were obtained from community-dwelling older adults aged 65 years and older in Arao City, Japan (baseline: 1,488; follow-up: 730). Serum ATX and dehydroepiandrosterone sulfate (DHEAS) levels were measured, and cognitive function and depressive symptoms were assessed using the Mini–Mental State Examination (MMSE) and Geriatric Depression Scale (GDS). Multiple linear regression models were applied to examine any cross-sectional associations among serum ATX levels, adrenal steroids, and MMSE/GDS scores. Longitudinal analyses assessed whether baseline serum ATX levels predicted 6-year changes in MMSE and GDS scores, with additional sex-stratified analyses and propensity score matching conducted to address any potential follow-up bias.

**Results:** After adjusting for age and sex, DHEAS levels were inversely associated with ATX levels. Cross-sectional analyses identified no association between serum ATX levels and MMSE/GDS scores. However, longitudinal analyses demonstrated significant associations between baseline serum ATX levels and changes in MMSE scores, particularly among women, and remained significant after multivariate and propensity score–matched analyses.

**Conclusion:** This study provides epidemiological evidence of an inverse association between DHEAS and ATX, indicating a potential link between adrenal endocrine function and ATX regulation. In older women, baseline serum ATX levels were associated with inter-individual variability in subsequent cognitive trajectories, indicating a potential role for ATX in age-related cognitive changes.

## Introduction

As the global population ages, the quality of life (QOL) of older adults has become a major focus of attention. Cognitive decline and depressive symptoms, both common among older adults, are considered neuropsychiatric dysfunctions that significantly impact QOL, and commonly co-occur and contribute to disability, loss of independence, and increased mortality among older adults. Due to overlapping symptoms and complex disease progression, diagnosis based solely on clinical presentation is extremely difficult in the early disease stages^1^. Increasing evidence suggests that cognitive and affective dysfunctions share similar neurobiological mechanisms^2^.

Prior studies have suggested that autotaxin (ATX) and its downstream lysophosphatidic acid (LPA) signaling are involved in neuropsychiatric disorders^3–5^. The primary physiological function of ATX, a secreted lysophospholipase D, is to catalyze the conversion of lysophosphatidylcholine to LPA^4,6^. Animal studies have shown that LPA receptor 1 (LPAR1)-deficient mice exhibit depression-like behavior and memory impairment^7, 8^. Our prior clinical study of patients with severe major depressive disorder (MDD) revealed that, patients with MDD had significantly lower ATX levels in both the serum and cerebrospinal fluid (CSF), as well as reduced levels of certain LPA species in the CSF compared with healthy controls^9, 10^. Collectively, these findings indicate that the ATX–LPA signaling pathway plays an important role in the regulation of affective and cognitive functions; however, population-based studies are required to further substantiate this hypothesis. Nevertheless, to date, no longitudinal cohort studies have examined whether changes in ATX levels are associated with subsequent alterations in cognitive function or depressive symptoms. ATX is a known biomarker of liver fibrosis, and is associated with other physiological factors, such as sex differences and obesity^11–13^. However, most studies on this marker have focused on middle-aged populations or clinical samples, and the determinants and physiological variability of ATX levels in community-dwelling older adults have not yet been well characterized.

Previous human and animal studies have shown that exogenous corticosteroid use alters ATX concentration. For example, serum ATX levels significantly decrease following oral prednisolone treatment in patients with autoimmune diseases^14^. In mouse models, dexamethasone reduced ATX activity and LPA levels in the plasma, brain, and adipose tissue^15^. However, the relationship between corticosteroids and ATX is limited to exogenous steroid use. To address this gap, we focused on cortisol and dehydroepiandrosterone sulfate (DHEAS), both endogenous adrenal steroids derived from the hypothalamic–pituitary–adrenal (HPA) axis^16^ closely associated with neuropsychiatric dysfunction. These hormones are clinically measurable and serve as markers of short-term reactivity and long-term physiological state^17^. Clarifying these associations may provide insights into the biological variability of ATX in later life, and its potential relevance to age-related neuropsychiatric outcomes.

This study aimed to conduct a comprehensive exploratory analysis incorporating factors previously indicated to be involved in ATX fluctuations among community-dwelling older adults in Japan, and to examine whether ATX levels are associated with cognitive and depressive symptoms using both cross-sectional and longitudinal data.

## Methods

### Study design and participants

This study was conducted within the framework of the Japan Prospective Studies Collaboration for Aging and Dementia (JPSC-AD), a nationwide prospective cohort study of dementia launched in 2016 across eight locations in Japan^18^. The target population comprised community-dwelling older adults aged 65 years residing in Japan. For the present analysis, we used data from the Arao cohort, one of the JPSC-AD study sites.

The baseline survey (first wave), encompassing 1,577 participants, was conducted between November 2016 and February 2017. A comprehensive follow-up survey was conducted from October 2022 to December 2023, approximately five to six years later.

Data for both the baseline and follow-up surveys were collected with the approval of the Ethics Committee of Kumamoto University (approval number: GENOME-333). Written informed consent was obtained from the participants or their legally authorized representatives. The present study analyzed the stored biological samples and associated data as secondary research, which was approved separately by the same committee (approval number: RINRI-2915).

In the cross-sectional analysis, 89 participants were excluded because of missing data or refusal to allow secondary data use, yielding a final cohort of 1,488 participants for the baseline analysis. For the longitudinal analysis, 758 participants were excluded because of missing follow-up data, leaving a final sample of 730 participants. The participant exclusion process is illustrated in Figure 1.

**Figure 1.**
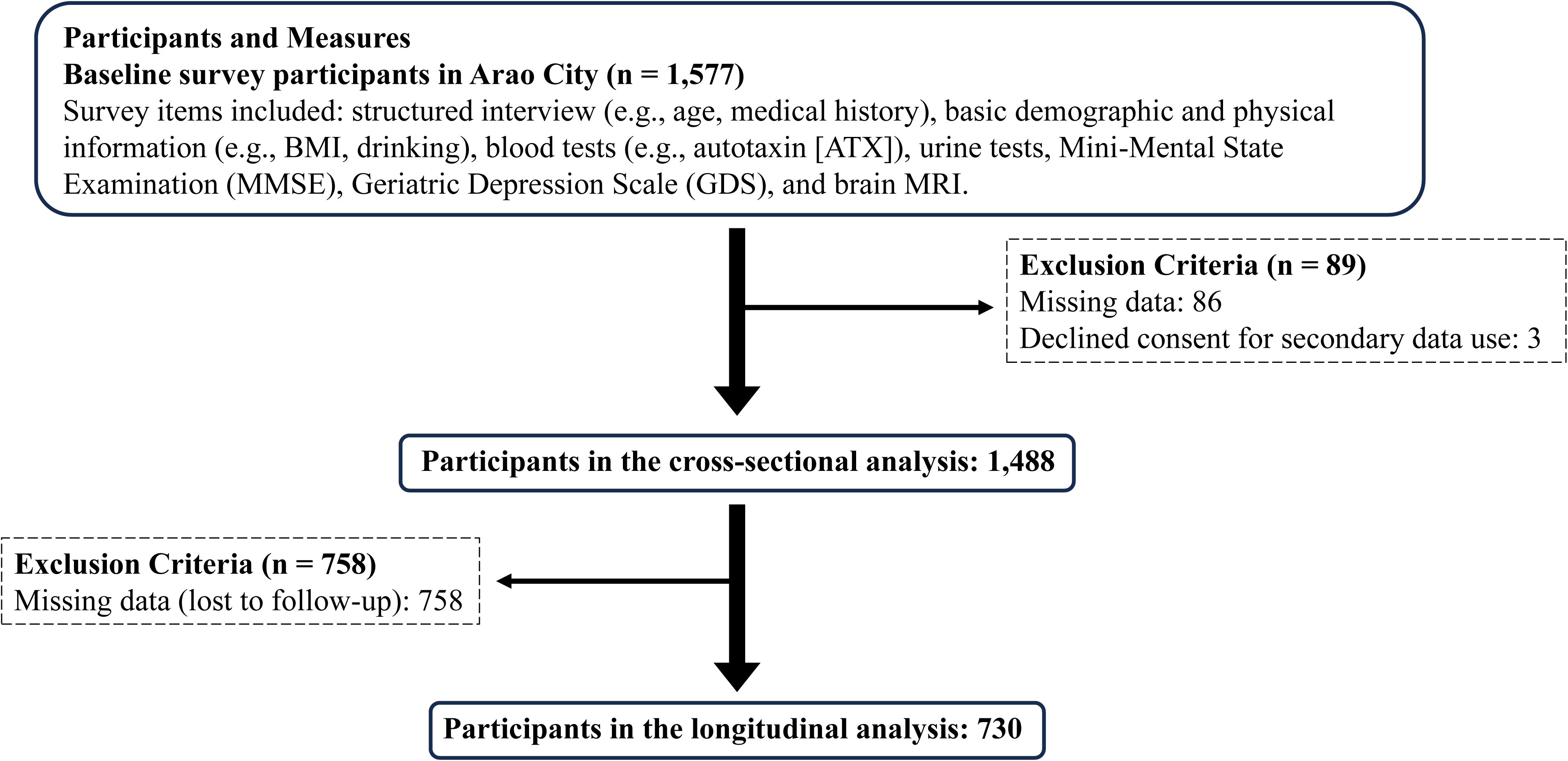
Flowchart showing participant enrolment. Of the 1,577 participants enrolled for the baseline survey in Arao City, 89 were excluded because of missing data or declining consent for secondary data use, leaving 1,488 participants for the cross-sectional analysis. Among them, 758 were lost to follow-up, leaving 730 for inclusion in the longitudinal analysis.

### Procedure

Standardized approaches for questionnaires, blood tests, and dementia diagnoses were applied across all study sites, as previously described^18^. Depressive symptoms were assessed using the Geriatric Depression Scale (GDS) short version,^19^ and cognitive function was assessed using the Japanese Test Form of the Mini-Mental State Examination (MMSE)^20^. Trained researchers administered the GDS and MMSE through interviews. Depression and dementia were clinically diagnosed according to the Diagnostic and Statistical Manual of Mental Disorders, Fourth Edition (DSM-IV)^21^ and Third Edition Revised (DSM-III-R)^22^, respectively.

Information was collected on the following covariates: sex, age, educational attainment, body mass index (BMI), daily physical activity, living status, smoking and alcohol consumption, history of stroke, head trauma, cardiovascular disease, and current medication use. Medical conditions were defined by the following factors: Hypertension, systolic blood pressure ≥140 mmHg or a diastolic blood pressure ≥90 mmHg; Diabetes mellitus, fasting blood glucose ≥126 mg/dL, a casual blood glucose ≥200 mg/dL, or an HbA1c ≥6.5%; Hypercholesterolemia, total serum cholesterol ≥220 mg/dL; Cardiac dysfunction, a history of atrial fibrillation, left ventricular hypertrophy, or arrhythmia; Renal dysfunction, abnormal estimated glomerular filtration rate (eGFR) <60 mL/min/1.73 m² and/or an elevated urine albumin-to-creatinine ratio ≥30 mg/g; Liver fibrosis, FIB-3 index ≥3.41^23^;; Inflammation, high-sensitivity C-reactive protein >0.3 mg/L; Thyroid dysfunction, thyroid-stimulating hormone level <0.5 μIU/mL or >4.5 μIU/mL, or a free thyroxine (T4) <0.8 ng/dL or >1.8 ng/dL; Obesity, BMI ≥25.

### Biomarker measurements

Blood samples were collected from participants, and serum was stored at −80°C until analysis. The serum concentrations of ATX, cortisol, and DHEAS were measured in stored baseline samples. Serum ATX, cortisol, and DHEAS concentrations were quantified using an AIA-CL 2400 automated chemiluminescent enzyme immunoassay analyzer (Tosoh Co., Tokyo, Japan), using the manufactured reagents. ATX was measured using sandwich immunoassay, while cortisol and DHEAS were measured using one-step delayed competitive immunoassays, performed according to the manufacturers’ instructions.

### Brain imaging analysis

Baseline participants in the JPSC-AD group underwent brain MRI at Arao Municipal Hospital using a 1.5-Tesla Philips Ingenia CX Dual Scanner (Philips Healthcare, Best, Netherlands) or Omuta Tenryo Hospital using a 1.5-Tesla GE Signa HDxt Ver.23 Scanner (GE Healthcare). The detailed scanner protocols have been described previously^24^. Volumetric segmentation was conducted using FreeSurfer version 5.3 (https://surfer.nmr.mgh.harvard.edu/) on CentOS 6, following standard preprocessing procedures^18^. Using the Desikan– Killany Atlas, we measured the absolute volumes of five cortical regions (the occipital lobe, temporal lobe, frontal lobe, parietal lobe, and insula), seven subcortical regions (the hippocampus, thalamus proper, accumbens area, amygdala, pallidum, caudate, and putamen), and the choroid plexus^25^. Each regional volume was normalized to the total intracranial volume. MRI analyses were conducted on the baseline participants. After sequential exclusion of those with missing data, stroke, head trauma, or dementia, 1,316 participants were included in the analysis. A detailed flow diagram is shown in Supplementary Figure 1.

### Statistical analysis

To examine univariate associations between serum ATX levels and participant characteristics, participants were stratified into sex-specific ATX quartiles. The Jonckheere-Terpstra trend test (for continuous variables) and Cochran-Armitage trend test (for categorical variables) were used to test for trends across ATX quartiles, treating quartiles as ordinal variables.

To identify any independent determinants of serum ATX levels, multivariate linear regression analyses were conducted using serum ATX as the dependent variable. Covariates included sex, age, liver fibrosis, DHEAS score, obesity, and corticosteroid use. Cortisol was examined in a separate model given its biological variability and correlation with DHEAS.

Cross-sectional and longitudinal analyses were conducted to examine the association between serum ATX levels and cognitive or depressive symptoms. Cross-sectional associations at baseline were examined using multiple linear regression models, with MMSE or GDS scores as dependent variables adjusted for age, sex, liver fibrosis, DHEAS score, obesity, and corticosteroid use.

The primary longitudinal analysis examined whether baseline serum ATX levels predicted 6-year changes in MMSE and GDS scores (ΔMMSE and ΔGDS). Changes were calculated as the follow-up score (2022) minus the baseline score (2016). Three sequential models were constructed: Model 1 was adjusted for baseline age, sex, baseline ATX level, and the corresponding baseline score; Model 2 was additionally adjusted for pre-specified ATX-related variables (liver fibrosis, DHEAS, obesity, and corticosteroid use); and Model 3 was further adjusted for established dementia-related covariates including education, depression, hypertension, diabetes, stroke history, living alone, drinking, smoking, and physical inactivity.

As a sensitivity analysis to mitigate any potential follow-up bias, propensity scores predicting follow-up participation were estimated using logistic regression, including all covariates in Model 3. One-to-one nearest-neighbor matching without replacement was performed, and regression analyses were repeated for the matched sample.

To examine associations between serum ATX levels and intracranial structural volumes, multiple linear regression models were constructed using the regional brain volumes (normalized to intracranial volume) as the dependent variables and serum ATX levels, age, sex, and MRI scanner type as covariates.

All statistical analyses were conducted using the JMP software (Student’s Edition 18). Statistical significance was set at p < 0.05. Benjamini-Hochberg false discovery rate (FDR) correction was applied for multiple comparisons as appropriate.

## Results

### Sample characteristics

At baseline, 1,488 participants were included in the cross-sectional analysis (904 females and 584 males). The baseline characteristics of the study participants are summarized in Table 1. The distribution of serum ATX levels according to sex is presented in Figure 2. Mean serum ATX levels were significantly higher in females (0.85 ± 0.23 ng/mL) than males (0.67 ± 0.17 ng/mL, p < 0.001). Serum ATX levels were divided into sex-specific quartiles (Q1–Q4), and distributions of demographic, lifestyle, and clinical variables were compared across the quartiles using trend tests. Higher ATX quartiles were consistently associated with older age and a greater prevalence of liver fibrosis and renal dysfunction in both sexes. Serum DHEAS levels decreased significantly with increasing ATX quartiles in both males and females. Among women, the proportion of participants living alone increased with increasing ATX quartiles. In contrast, among men, the MMSE score and proportion of participants with more than nine years of education decreased as ATX quartiles increased. Other demographic, lifestyle, and clinical variables were not significantly associated with the ATX quartiles after FDR adjustment.

**Figure 2.**
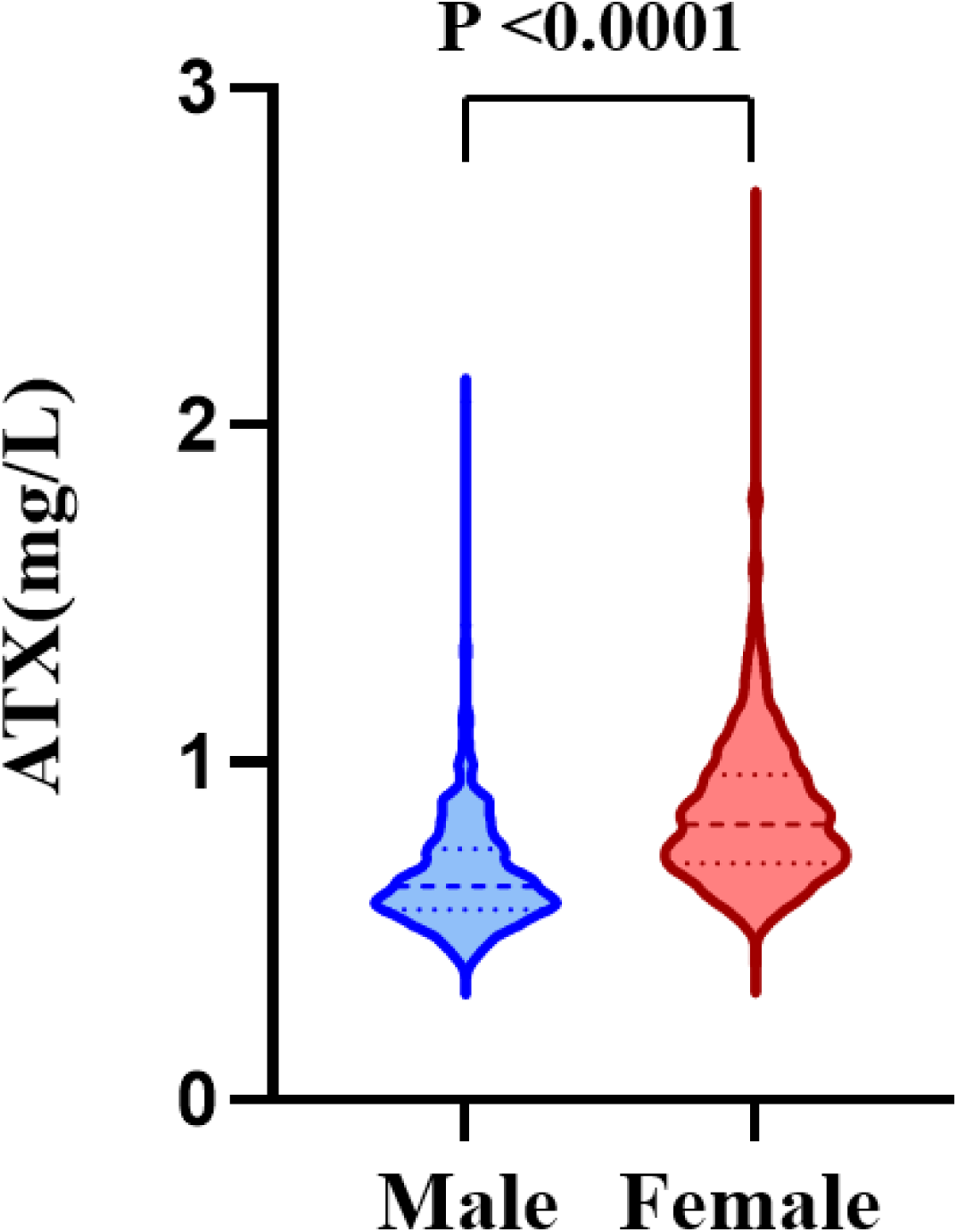
Distribution of serum ATX levels by sex at baseline. Violin plots showing the distribution of serum autotaxin (ATX) levels in male and female participants. The central line indicates the median and the dotted lines represent the interquartile range. Serum ATX levels were significantly higher in females than males (P < 0.0001).

**Table 1.**
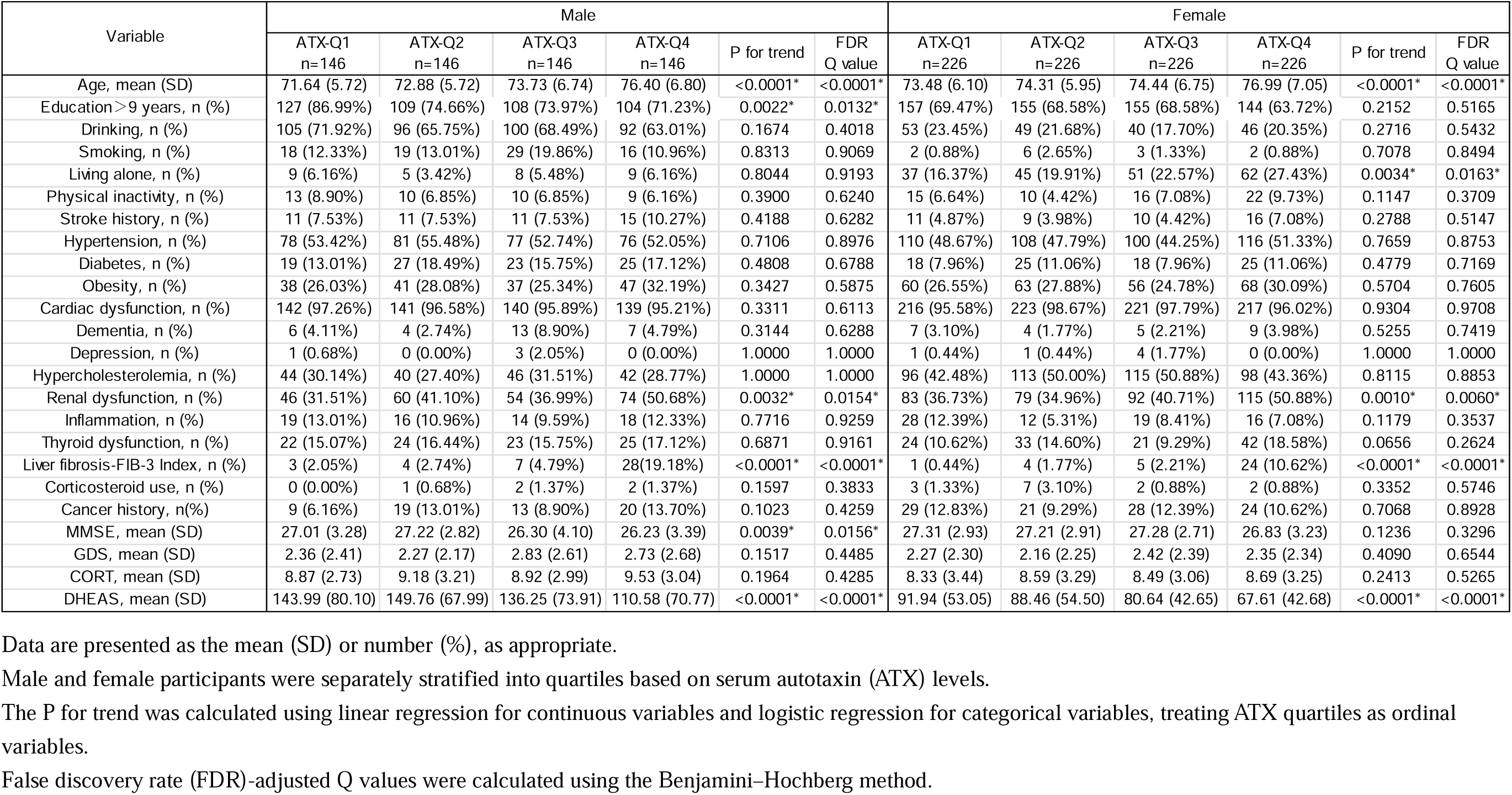
Baseline characteristics of participants stratified by serum ATX quartiles and sex.

| Variable | Male |  |  |  |  |  | Female |  |  |  |  |  |
| --- | --- | --- | --- | --- | --- | --- | --- | --- | --- | --- | --- | --- |
|  | ATX-Q1<br>n=146 | ATX-Q2<br>n=146 | ATX-Q3<br>n=146 | ATX-Q4<br>n=146 | P for trend | FDR<br>Q value | ATX-Q1<br>n=226 | ATX-Q2<br>n=226 | ATX-Q3<br>n=226 | ATX-Q4<br>n=226 | P for trend | FDR<br>Q value |
| Age, mean (SD) | 71.64 (5.72) | 72.88 (5.72) | 73.73 (6.74) | 76.40 (6.80) | <0.0001* | <0.0001* | 73.48 (6.10) | 74.31 (5.95) | 74.44 (6.75) | 76.99 (7.05) | <0.0001* | <0.0001* |
| Education > 9 years, n (%) | 127 (86.99%) | 109 (74.66%) | 108 (73.97%) | 104 (71.23%) | 0.0022* | 0.0132* | 157 (69.47%) | 155 (68.58%) | 155 (68.58%) | 144 (63.72%) | 0.2152 | 0.5165 |
| Drinking, n (%) | 105 (71.92%) | 96 (65.75%) | 100 (68.49%) | 92 (63.01%) | 0.1674 | 0.4018 | 53 (23.45%) | 49 (21.68%) | 40 (17.70%) | 46 (20.35%) | 0.2716 | 0.5432 |
| Smoking, n (%) | 18 (12.33%) | 19 (13.01%) | 29 (19.86%) | 16 (10.96%) | 0.8313 | 0.9069 | 2 (0.88%) | 6 (2.65%) | 3 (1.33%) | 2 (0.88%) | 0.7078 | 0.8494 |
| Living alone, n (%) | 9 (6.16%) | 5 (3.42%) | 8 (5.48%) | 9 (6.16%) | 0.8044 | 0.9193 | 37 (16.37%) | 45 (19.91%) | 51 (22.57%) | 62 (27.43%) | 0.0034* | 0.0163* |
| Physical inactivity, n (%) | 13 (8.90%) | 10 (6.85%) | 10 (6.85%) | 9 (6.16%) | 0.3900 | 0.6240 | 15 (6.64%) | 10 (4.42%) | 16 (7.08%) | 22 (9.73%) | 0.1147 | 0.3709 |
| Stroke history, n (%) | 11 (7.53%) | 11 (7.53%) | 11 (7.53%) | 15 (10.27%) | 0.4188 | 0.6282 | 11 (4.87%) | 9 (3.98%) | 10 (4.42%) | 16 (7.08%) | 0.2788 | 0.5147 |
| Hypertension, n (%) | 78 (53.42%) | 81 (55.48%) | 77 (52.74%) | 76 (52.05%) | 0.7106 | 0.8976 | 110 (48.67%) | 108 (47.79%) | 100 (44.25%) | 116 (51.33%) | 0.7659 | 0.8753 |
| Diabetes, n (%) | 19 (13.01%) | 27 (18.49%) | 23 (15.75%) | 25 (17.12%) | 0.4808 | 0.6788 | 18 (7.96%) | 25 (11.06%) | 18 (7.96%) | 25 (11.06%) | 0.4779 | 0.7169 |
| Obesity, n (%) | 38 (26.03%) | 41 (28.08%) | 37 (25.34%) | 47 (32.19%) | 0.3427 | 0.5875 | 60 (26.55%) | 63 (27.88%) | 56 (24.78%) | 68 (30.09%) | 0.5704 | 0.7605 |
| Cardiac dysfunction, n (%) | 142 (97.26%) | 141 (96.58%) | 140 (95.89%) | 139 (95.21%) | 0.3311 | 0.6113 | 216 (95.58%) | 223 (98.67%) | 221 (97.79%) | 217 (96.02%) | 0.9304 | 0.9708 |
| Dementia, n (%) | 6 (4.11%) | 4 (2.74%) | 13 (8.90%) | 7 (4.79%) | 0.3144 | 0.6288 | 7 (3.10%) | 4 (1.77%) | 5 (2.21%) | 9 (3.98%) | 0.5255 | 0.7419 |
| Depression, n (%) | 1 (0.68%) | 0 (0.00%) | 3 (2.05%) | 0 (0.00%) | 1.0000 | 1.0000 | 1 (0.44%) | 1 (0.44%) | 4 (1.77%) | 0 (0.00%) | 1.0000 | 1.0000 |
| Hypercholesterolemia, n (%) | 44 (30.14%) | 40 (27.40%) | 46 (31.51%) | 42 (28.77%) | 1.0000 | 1.0000 | 96 (42.48%) | 113 (50.00%) | 115 (50.88%) | 98 (43.36%) | 0.8115 | 0.8853 |
| Renal dysfunction, n (%) | 46 (31.51%) | 60 (41.10%) | 54 (36.99%) | 74 (50.68%) | 0.0032* | 0.0154* | 83 (36.73%) | 79 (34.96%) | 92 (40.71%) | 115 (50.88%) | 0.0010* | 0.0060* |
| Inflammation, n (%) | 19 (13.01%) | 16 (10.96%) | 14 (9.59%) | 18 (12.33%) | 0.7716 | 0.9259 | 28 (12.39%) | 12 (5.31%) | 19 (8.41%) | 16 (7.08%) | 0.1179 | 0.3537 |
| Thyroid dysfunction, n (%) | 22 (15.07%) | 24 (16.44%) | 23 (15.75%) | 25 (17.12%) | 0.6871 | 0.9161 | 24 (10.62%) | 33 (14.60%) | 21 (9.29%) | 42 (18.58%) | 0.0656 | 0.2624 |
| Liver fibrosis-FIB-3 Index, n (%) | 3 (2.05%) | 4 (2.74%) | 7 (4.79%) | 28 (19.18%) | <0.0001* | <0.0001* | 1 (0.44%) | 4 (1.77%) | 5 (2.21%) | 24 (10.62%) | <0.0001* | <0.0001* |
| Corticosteroid use, n (%) | 0 (0.00%) | 1 (0.68%) | 2 (1.37%) | 2 (1.37%) | 0.1597 | 0.3833 | 3 (1.33%) | 7 (3.10%) | 2 (0.88%) | 2 (0.88%) | 0.3352 | 0.5746 |
| Cancer history, n(%) | 9 (6.16%) | 19 (13.01%) | 13 (8.90%) | 20 (13.70%) | 0.1023 | 0.4259 | 29 (12.83%) | 21 (9.29%) | 28 (12.39%) | 24 (10.62%) | 0.7068 | 0.8928 |
| MMSE, mean (SD) | 27.01 (3.28) | 27.22 (2.82) | 26.30 (4.10) | 26.23 (3.39) | 0.0039* | 0.0156* | 27.31 (2.93) | 27.21 (2.91) | 27.28 (2.71) | 26.83 (3.23) | 0.1236 | 0.3296 |
| GDS, mean (SD) | 2.36 (2.41) | 2.27 (2.17) | 2.83 (2.61) | 2.73 (2.68) | 0.1517 | 0.4485 | 2.27 (2.30) | 2.16 (2.25) | 2.42 (2.39) | 2.35 (2.34) | 0.4090 | 0.6544 |
| CORT, mean (SD) | 8.87 (2.73) | 9.18 (3.21) | 8.92 (2.99) | 9.53 (3.04) | 0.1964 | 0.4285 | 8.33 (3.44) | 8.59 (3.29) | 8.49 (3.06) | 8.69 (3.25) | 0.2413 | 0.5265 |
| DHEAS, mean (SD) | 143.99 (80.10) | 149.76 (67.99) | 136.25 (73.91) | 110.58 (70.77) | <0.0001* | <0.0001* | 91.94 (53.05) | 88.46 (54.50) | 80.64 (42.65) | 67.61 (42.68) | <0.0001* | <0.0001* |

Data are presented as the mean (SD) or number (%), as appropriate.
Male and female participants were separately stratified into quartiles based on serum autotaxin (ATX) levels.
The P for trend was calculated using linear regression for continuous variables and logistic regression for categorical variables, treating ATX quartiles as ordinal variables.
False discovery rate (FDR)-adjusted Q values were calculated using the Benjamini–Hochberg method.

### Associations between serum ATX levels and endogenous adrenal steroids

To further clarify whether endogenous adrenal steroid levels (cortisol and DHEAS) are associated with serum ATX levels, independent of other known determinants, we examined the relationships among serum cortisol, DHEAS, and ATX levels, while considering the differences in biological stability between these two hormones.

Cortisol exhibits a pronounced circadian rhythm, and is strongly influenced by sampling-related conditions^26^. Therefore, we first analyzed the associations of blood sampling time, presampling exercise, and fasting status with cortisol and DHEAS levels. As presented in Supplementary Table 1, cortisol levels were significantly associated with sampling time, pre-sampling exercise, and fasting, while DHEAS levels were unaffected by these factors. Despite these differences in sampling-related variability, serum cortisol and DHEAS levels showed a significant positive correlation, as assessed by Pearson’s correlation coefficient (r = 0.15, p < 0.001).

We conducted multiple linear regression analyses to further examine the associations between serum cortisol, DHEAS, and ATX levels. Based on prior evidence, serum ATX levels are influenced by multiple factors including sex, liver fibrosis, age, and obesity^11–13, 27^. The results revealed that, in addition to sex, age, liver fibrosis, and obesity, serum DHEAS levels were significantly associated with serum ATX levels (β = −0.0005, p < 0.0001) (Table 2). In contrast, cortisol levels showed no significant association with serum ATX levels after adjusting for relevant sampling-related variables (p = 0.0692) (Supplementary Table 2).

**Table 2.** Multivariate linear regression analysis of factors associated with serum ATX levels.

| Variable | $\beta$ (95% CI) | p-value |
| --- | --- | --- |
| Sex (Female) | 0.081 (0.070~0.092) | <0.0001* |
| Age | 0.005 (0.003~0.006) | <0.0001* |
| Liver fibrosis-FIB-3 Index (Yes) | 0.124 (0.102~0.147) | <0.0001* |
| DHEAS | -0.0005 (-0.0007~-0.0003) | <0.0001* |
| Obesity (Yes) | 0.012 (0.001~0.023) | 0.0303* |
| Corticosteroid use (Yes) | -0.012 (-0.055~0.032) | 0.5939 |

Multivariate linear regression was conducted with the serum ATX level (mg/L) as the dependent variable (n = 1,488).

### Cross-sectional association of serum ATX with cognitive function and depressive symptoms

Next, we examined whether serum ATX levels were associated with cognitive function or depressive symptoms at baseline. Multiple linear regression analyses were conducted using MMSE and GDS scores as dependent variables, adjusting for age, sex, liver fibrosis, DHEAS score, obesity, and corticosteroid use (Table 3). Serum ATX levels were not associated with the MMSE scores in either sex. However, higher serum ATX levels were associated with higher GDS scores in males (β = 1.455, p = 0.035), while no association was observed in females.

**Table 3.** Sex-stratified multivariate linear regression analysis of baseline MMSE and GDS scores.

| Dependent variable | Sample | $\beta$ for serum ATX<br>(95% CI) | p-value |
| --- | --- | --- | --- |
| GDS | all | 0.191(-0.441~0.824) | 0.5528 |
|  | male | 1.455(0.106~2.805) | 0.0346* |
|  | female | -0.188(-0.896~0.519) | 0.6012 |
| MMSE | all | 0.619(-0.154~1.392) | 0.1163 |
|  | male | 0.842(-0.895~2.579) | 0.3416 |
|  | female | 0.492(-0.338~1.321) | 0.2452 |

Multivariate linear regression analyses were conducted using the baseline MMSE or GDS score as the dependent variable.
$\beta$ represents the unstandardized regression coefficient for serum ATX level.
All models were adjusted for age, sex (except sex-stratified analyses), liver fibrosis, DHEAS, obesity, and corticosteroid use.
Male, n = 584; Female, n = 904.

### Cross-sectional analyses using follow-up data

To evaluate whether the cross-sectional associations observed at baseline remained consistent over time, the analyses were repeated using serum ATX levels, serum DHEAS levels, MMSE scores, and GDS scores obtained from 675 participants who completed the 2022 follow-up assessment, after excluding 55 participants with missing data at follow-up. Consistent with baseline findings, serum DHEAS levels remained significantly associated with serum ATX levels (Supplementary Table 3). Similarly, no significant association was observed between serum ATX levels and MMSE scores in either sex at the follow-up (Supplementary Table 4). In contrast, no association between serum ATX levels and GDS scores in males was observed in the follow-up data.

### Associations between serum ATX levels and brain regional volumes

Given the known role of ATX–LPA signaling in the central nervous system, we further examined whether serum ATX levels were associated with regional brain volumes. Associations between serum ATX levels and regional brain volumes were evaluated using multiple linear regression analyses adjusted for age, sex, and scanner type (Supplementary Table 5). In uncorrected analyses, a nominal association was identified between ATX and choroid plexus volume (β = 0.012, p = 0.0334). However, this association lost significance after correcting for multiple comparisons (q = 0.4342), and no other regional volumes showed significant associations with ATX.

### Longitudinal association of serum ATX with cognitive function and depressive symptoms

Longitudinal changes in MMSE and GDS scores were analyzed among the 730 participants who completed the 2022 follow-up (Table 4). In the fully adjusted model (Model 3), baseline serum ATX levels were found to be significantly associated with changes in MMSE scores (β = 1.661, p = 0.0033). This association was consistent across all three statistical models. In contrast, baseline serum ATX levels were not found to be associated with longitudinal changes in the GDS scores. The sex-stratified analyses indicated that this association was primarily observed in women. In females, serum ATX levels were significantly associated with changes in MMSE scores across all three models, whereas no significant associations were observed in males.

**Table 4.**
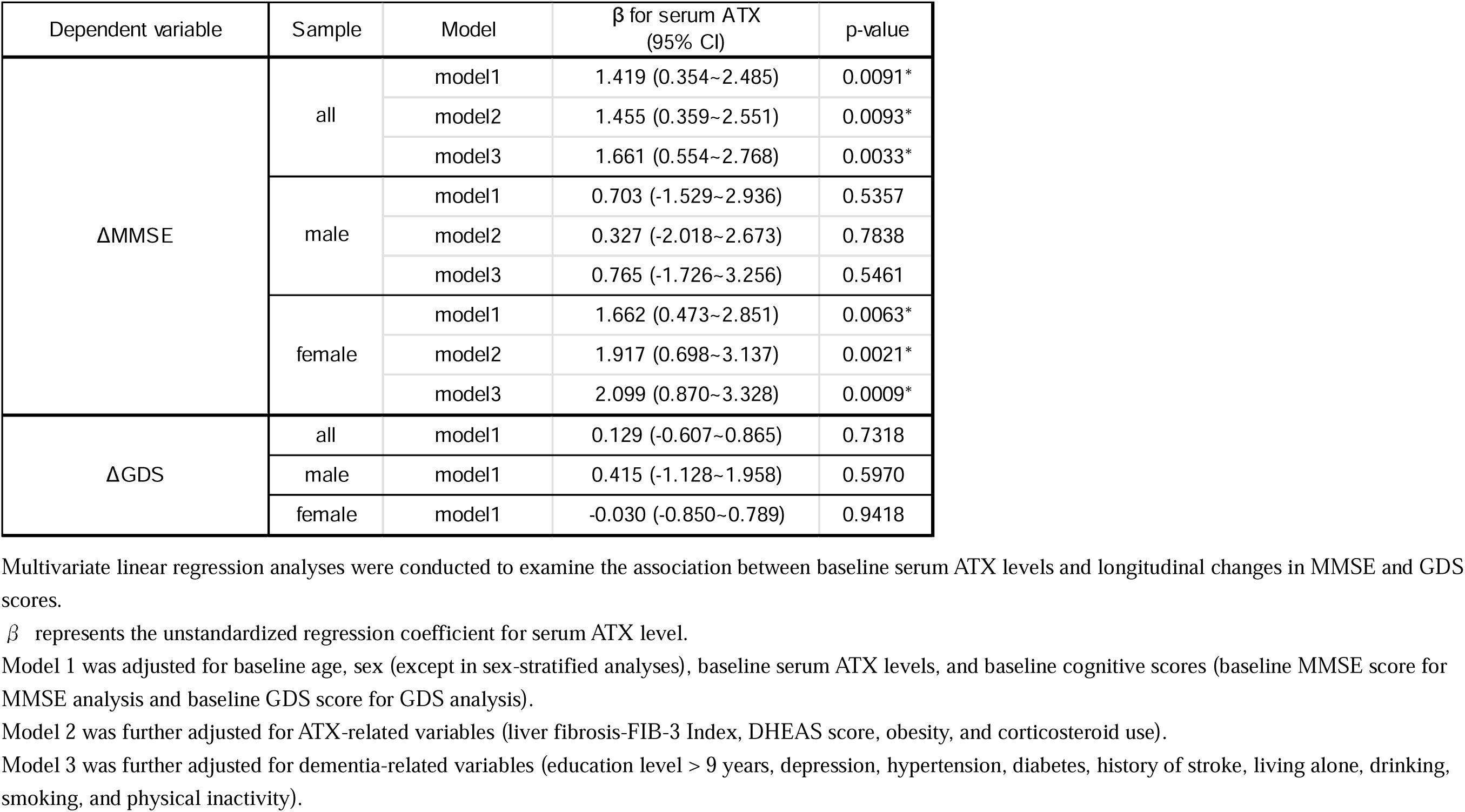
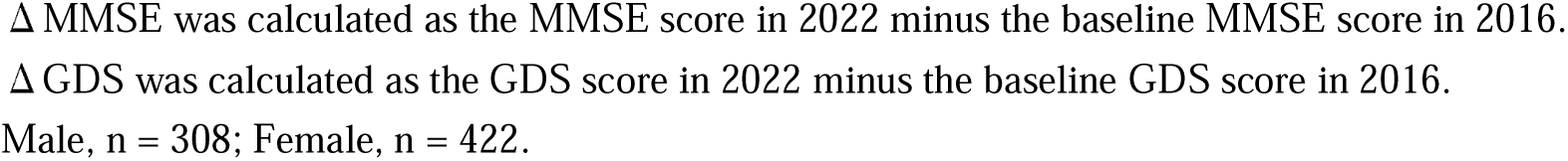
Results of multivariate linear regression for changes in MMSE and GDS scores.

### Assessment of follow-up bias and propensity score matching analysis

Baseline characteristics were compared between participants who completed follow-up and those lost to follow-up (Supplementary Table 6). Statistically significant differences in several demographic and clinical variables were observed between the two groups, suggesting a potential follow-up bias.

To mitigate this bias, propensity score matching was conducted based on the covariates included in Model 3 (Table 4). After matching, the total sample size decreased from 1,488 to 994, with 497 participants retained in each group (Supplementary Table 6). Using the matched dataset, multiple linear regression analysis was repeated to examine the factors associated with changes in MMSE scores (Supplementary Table 7). The association between serum ATX levels and longitudinal changes in cognitive function remained robust even after propensity score matching.

## Discussion

In this community-based cohort of older adults, we examined the determinants of serum ATX levels and their associations with cognitive function and depressive symptoms. Two principal findings emerged. First, serum ATX levels were independently associated with demographic and metabolic factors, including age, sex, liver fibrosis, obesity, and circulating DHEAS levels. Second, baseline serum ATX levels were modestly but consistently associated with longitudinal changes in cognitive performance over approximately 6 years, particularly in women. These findings extend previous clinical observations and suggest that circulating ATX may reflect biological variability relevant to cognitive aging.

### I. Determinants of serum ATX levels in older adults

Our findings confirmed several previously reported determinants of serum ATX levels, including sex, liver fibrosis, and obesity^11–13, 28^. In addition, serum ATX levels were positively associated with age in this cohort of community-dwelling older adults. This finding differs from a previous study conducted mainly in middle-aged adults, which reported an inverse association between ATX and age in men and no clear association in women^28^. The discrepancy may reflect differences in age distribution between study populations, and suggests that the physiological correlates of ATX may differ in later life.

A novel finding of this study was the robust inverse association between serum DHEAS and ATX levels. This relationship was observed at baseline and in the follow-up cross-sectional analysis, supporting its consistency. Although exogenous glucocorticoids have been reported to reduce ATX levels^14^, evidence regarding endogenous adrenal steroids has been limited. In our cohort, DHEAS showed a stable and independent association with ATX, whereas cortisol did not retain statistical significance. One possible explanation is that serum cortisol was strongly influenced by blood sampling conditions in this study. Residual confounding related to sampling conditions may therefore have attenuated its association with ATX. Although cortisol and DHEAS have distinct biological roles,^16, 17^ their positive correlation in this cohort suggests that both partly reflect shared adrenal or HPA axis-related activity. Given the cross-sectional nature of this analysis, the directionality and mechanisms underlying this association cannot be determined. Nevertheless, these findings suggest that adrenal endocrine function may represent one component of the physiological variability of circulating ATX.

We further examined the association between serum ATX levels and regional brain volumes using MRI data. After correcting for multiple comparisons, no brain region showed a statistically significant association with serum ATX levels. In uncorrected analyses, a nominal association was identified between serum ATX levels and choroid plexus volume. Given that ATX expression is particularly high in the choroid plexus compared to other brain regions^29, 30^, this observation may reflect region-specific biological relevance, although the lack of significance after correction precludes any definitive conclusion.

### II. Clinical relevance of serum ATX in older adults

In contrast to our previous clinical studies in patients with severe MDD^9^, serum ATX levels were not consistently associated with depressive symptoms in this community-dwelling sample. This discrepancy may reflect important differences between clinically diagnosed MDD and subthreshold depressive symptoms in the general population. In addition, only a small number of participants met diagnostic criteria for MDD, and GDS is a self-reported measure that may be less reliable in individuals with cognitive impairment^31^. These findings suggest that ATX alterations observed in clinical depression may not be readily detectable among community-dwelling older adults with mostly subthreshold depressive symptoms.

The most clinically relevant finding of this study was that baseline serum ATX levels were associated with subsequent change in cognitive function over approximately 6 years. This association was robust across sequential adjustment models and remained significant after propensity score matching, supporting the possibility that serum ATX reflects biologically meaningful variation related to cognitive aging. Notably, no clear association was observed between ATX and MMSE scores in the cross-sectional analyses at either baseline or follow-up. This pattern may indicate that serum ATX is not simply a marker of current cognitive status, but rather is associated with inter-individual differences in cognitive trajectories over time.

The mechanisms underlying this association remain uncertain. Because ATX-LPA signaling has been implicated in affective and cognitive regulation in experimental studies^3, 4, 32^, one possible interpretation is that peripheral ATX may reflect biological processes relevant to brain aging and cognitive change. The inverse association between DHEAS and ATX further raises the possibility that endocrine factors related to HPA axis function contribute to these individual differences. However, the present observational data do not allow any causal inference. Further longitudinal studies incorporating repeated measurements of DHEAS, ATX, and cognitive function will be necessary to clarify these temporal and biological relationships.

The association between ATX and longitudinal cognitive change was more evident in women. Women also showed higher serum ATX levels than men, consistent with previous reports^28^, and higher circulating LPA levels have also been reported in women^33^. Although the basis of this sex difference remains unclear, sex-dependent behavioral effects related to ATX-LPA signaling have been reported in experimental studies^34, 35^. In addition, postmortem studies have reported reduced expression of ATX-related molecules, including ATX and LPAR1, in disorders such as MDD and Alzheimer’s disease^36–38^, supporting the potential relevance of this pathway to brain dysfunction. However, whether the clinical significance of ATX-LPA-related alterations differs by sex remains unknown. Future studies investigating the functional significance of the ATX-LPA pathway should therefore consider sex-informed analyses.

Several limitations should be noted. First, the longitudinal analysis included only participants who completed both baseline and follow-up assessments, and those who developed more severe neuropsychiatric impairment may have been less likely to attend follow-up, potentially resulting in a healthier analytic sample. Although we attempted to address this issue using propensity score matching, residual follow-up bias cannot be excluded. Second, incident dementia was not examined in this study. Longer follow-up with more comprehensive outcome ascertainment will be needed to determine whether serum ATX is associated with future dementia risk. Finally, the observational design precludes any conclusion regarding causality.

Despite these limitations, this study provides new epidemiological evidence that serum ATX levels in older adults are associated not only with known physiological factors but also with endogenous adrenal function, as reflected by DHEAS. In addition, baseline serum ATX levels were associated with subsequent cognitive change, particularly in women. These findings suggest that serum ATX may serve as a candidate biomarker related to inter-individual variability in cognitive trajectories in later life.

## Data Availability

All data produced in the present study are available upon reasonable request to the authors

## Acknowledgments

The authors would like to thank the residents of Arao City for participating in this study. We further gratefully acknowledge all staff members involved in this research and field surveys from the Division of Health, Arao City; Department of Neuropsychiatry, Kumamoto University; and various departments of the Faculty of Life Sciences, Kumamoto University; Arao Medical Association; and Ariake Medical Center (formerly Arao City Hospital). We wish to personally thank Drs. Saruwatari, Oniki, and Morita (Kumamoto University) for their valuable comments and constructive advice. The authors also thank the TOSOH Corporation for technical assistance with the measurement of serum biomarkers.

During the preparation of this manuscript, the authors used ChatGPT (GPT-5.2, OpenAI) and Editage’s English proofreading service to enhance the readability and proofread the English text. After using these services, the authors reviewed and edited the manuscript as necessary and take full responsibility for the content of this publication.

## Funding

This study was supported in part by the Japan Agency for Medical Research and Development (AMED; JP25dk0207053), and by a collaborative research grant from TOSOH Corporation.

## Disclosure statement

MT received collaborative research funding from TOSOH Corporation, which provided technical support for the measurement of serum ATX and DHEAS levels. The funder had no role in the study design, data analysis, data interpretation, or manuscript preparation. All other authors declare no competing interests.

## Author contributions

NK and MT conceived and designed the study. SS performed data analysis. NK, KY, MM and MT interpreted the data. SS and NK drafted the manuscript. All authors critically revised the manuscript and approved the final version.

## Figure legends

**Supplementary Figure 1.**
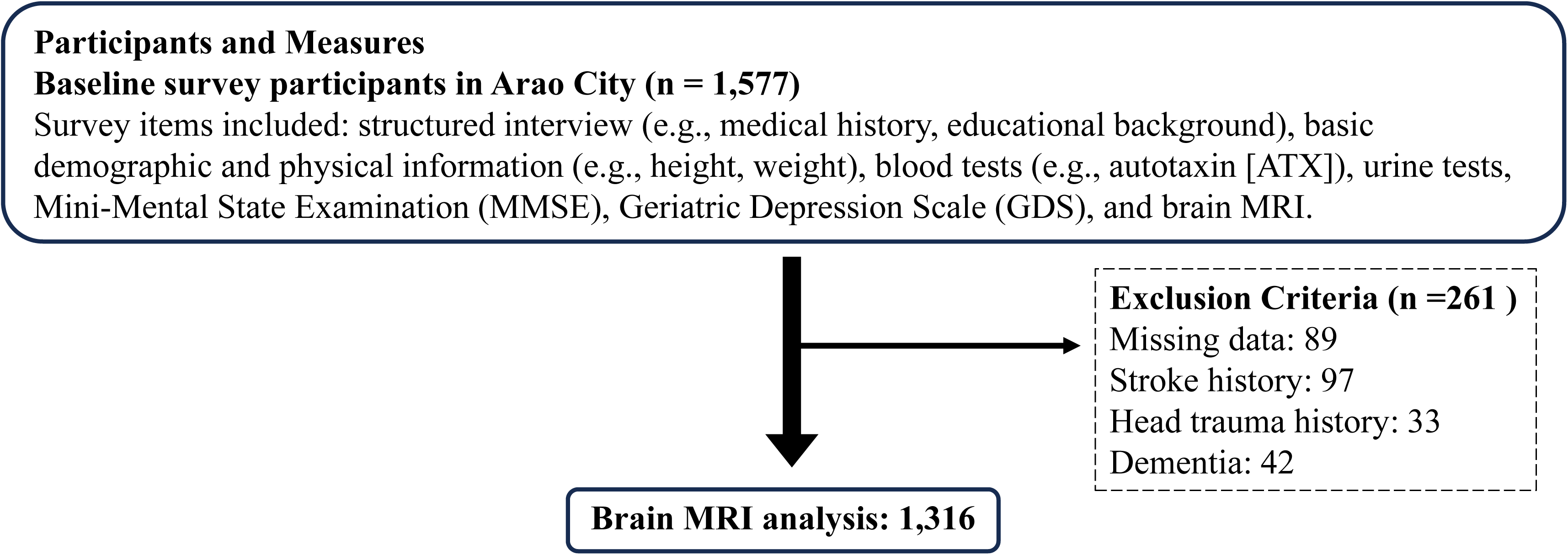
Flowchart of participant selection for brain MRI analysis. Baseline participants in the Arao City cohort underwent brain MRI examinations. Of the 1,577 participants enrolled at baseline, individuals were excluded due to missing data (n = 89), history of stroke (n = 97), head trauma (n = 33), or dementia (n = 42). These conditions were excluded because they may cause structural brain changes that could confound volumetric analyses. After these exclusions, a total of 1,316 participants were included in the brain MRI analysis.

**Supplementary Table 1.** Associations between sampling-related factors and cortisol/DHEAS levels.

|  | Cortisol |  | DHEA-S |  |
| --- | --- | --- | --- | --- |
| | $\beta$ (95% CI) | p value | $\beta$ (95% CI) | p value |
| Age | 0.0428 (0.018~0.067) | 0.0006* | -2.277 (-2.740~-1.815) | <0.0001* |
| Sex (Female) | -0.292 (-0.454~-0.129) | 0.0005* | -25.277 (-28.364~-22.190) | <0.0001* |
| Blood sampling time (AM) | -0.531 (-0.695~-0.368) | <0.0001* | -0.237 (-3.328~2.854) | 0.8806 |
| Fasting blood sample (Yes) | -0.248 (-0.493~-0.004) | 0.0466* | -2.053 (-6.686~2.580) | 0.3848 |
| Pre-sampling exercise (Yes) | 0.230 (0.0318~0.428) | 0.0230* | 1.952 (-1.802~5.707) | 0.3079 |

Multivariate linear regression analyses were conducted to examine the associations between sampling-related factors, cortisol, and DHEAS levels.
Blood sampling time was coded as AM vs PM.
Pre-sampling exercise (Yes) indicated exercise within 1 h before blood sampling.
n = 1,469.

**Supplementary Table 2.** Results of multivariate linear regression analysis of factors associated with serum ATX levels after adjusting for cortisol.

| Variable | $\beta$ (95% CI) | p value |
| --- | --- | --- |
| Sex (Female) | 0.093 (0.083~0.104) | <0.0001* |
| Age | 0.006 (0.004~0.007) | <0.0001* |
| Liver fibrosis-FIB-3 Index (Yes) | 0.121 (0.099~0.144) | <0.0001* |
| Cortisol | 0.003 (-0.0002~0.006) | 0.0692 |
| Blood sampling time (AM) | -0.010 (-0.020~0.001) | 0.0668 |
| Fasting blood sample (Yes) | -0.008 (-0.020~0.005) | 0.2230 |
| Pre-sampling exercise (Yes) | -0.005 (-0.020~0.011) | 0.5514 |
| Obesity (Yes) | 0.013 (0.002~0.024) | 0.0204* |
| Corticosteroid use (Yes) | 0.009 (-0.035~0.053) | 0.6841 |

Multivariate linear regression analysis was performed to examine factors associated with serum ATX levels after adjusting for cortisol levels.
Blood sampling time was coded as AM vs PM.
Pre-sampling exercise (Yes) indicated exercise within 1 h before blood sampling.
n = 1,469.

**Supplementary Table 3.** Multivariate linear regression analysis of factors associated with serum ATX levels at the 2022 follow-up.

| Variable | $\beta$ (95% CI) | p-value |
| --- | --- | --- |
| Sex (Female) | 0.074 (0.048~0.100) | <.0001* |
| Age | 0.005 (0.0003~0.010) | 0.0374* |
| Liver fibrosis-FIB-3 Index (Yes) | 0.215 (0.155~0.274) | <.0001* |
| DHEAS | -0.001 (-0.001~-0.0001) | 0.0126* |
| Obesity (Yes) | 0.034 (0.006~0.062) | 0.0187* |

Multivariate linear regression analysis was conducted to examine the factors associated with serum ATX levels at the 2022 follow-up. n = 675.

**Supplementary Table 4.** Sex-stratified multivariate linear regression analyses of MMSE and GDS scores at the 2022 follow-up.

| Dependent variable | Sample | $\beta$ for serum ATX (95% CI) | p-value |
| --- | --- | --- | --- |
| GDS | all | -0.233 (-0.735~0.269) | 0.3628 |
|  | male | -0.153 (-0.775~0.469) | 0.6287 |
|  | female | -0.486 (-1.446~0.474) | 0.3201 |
| MMSE | all | 0.451 (-0.203~1.106) | 0.1761 |
|  | male | 0.287 (-0.542~1.115) | 0.4965 |
|  | female | 1.028 (-0.197~2.253) | 0.0998 |

Multivariate linear regression analyses were conducted with MMSE or GDS scores at the 2022 follow-up as the dependent variable.
$\beta$ represents the unstandardized regression coefficient for serum ATX level.
All models were adjusted for age, sex (except sex-stratified analyses), liver fibrosis-FIB-3 Index, DHEA-S, and obesity.
Males, n = 286; Females, n = 389.

**Supplementary Table 5.** Multivariate linear regression analyses of associations between serum ATX levels and regional brain volumes.

| Brain region | $\beta$ for serum ATX (95% CI) | p value | q value |
| --- | --- | --- | --- |
| Choroid plexus | 0.012 (0.001~0.022) | 0.0334* | 0.4342 |
| Pallidum | -0.006 (-0.012~0.001) | 0.0853 | 0.5545 |
| Hippocampus | 0.010 (-0.005~0.026) | 0.1836 | 0.7956 |
| Putamen | -0.011 (-0.034~0.0116) | 0.3313 | 0.8051 |
| Amygdala | -0.003 (-0.008~0.003) | 0.3756 | 0.8051 |
| Parietal lobe | -0.062 (-0.200~0.076) | 0.3803 | 0.8051 |
| Caudate | -0.007 (-0.026~0.011) | 0.4335 | 0.8051 |
| Frontal lobe | -0.044 (-0.230~0.142) | 0.6410 | 0.8216 |
| Thalamus proper | 0.004 (-0.015~0.023) | 0.6856 | 0.8216 |
| Accumbens area | 0.0004 (-0.002~0.003) | 0.7697 | 0.8216 |
| Occipital lobe | 0.007 (-0.051~0.065) | 0.8184 | 0.8216 |
| Temporal lobe | -0.013 (-0.124~0.098) | 0.8190 | 0.8216 |
| Insula | -0.002 (-0.022~0.018) | 0.8216 | 0.8216 |

Multivariate linear regression analyses were conducted to examine the association between baseline serum ATX levels and the regional brain volume.
Regional brain volumes were normalized to intracranial volume and expressed as percentages of intracranial volume (%ICV).
$\beta$ represents the percentage-point difference in regional brain volume per 1-unit increase in serum ATX level (ng/mL).
All models were adjusted for age, sex, and MRI scanner type.
Q values were calculated using the Benjamini–Hochberg false discovery rate correction.

**Supplementary Table 6.**
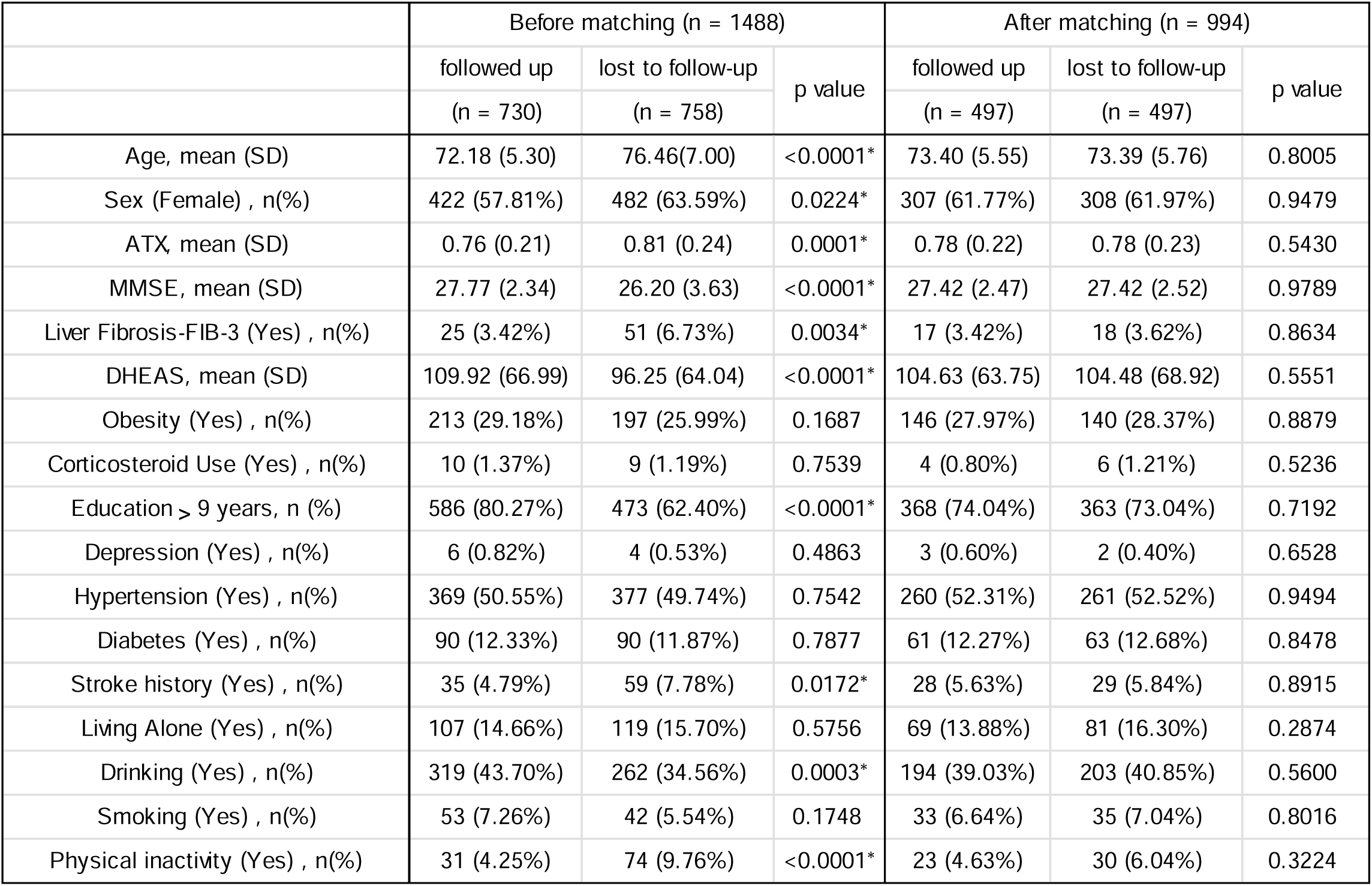
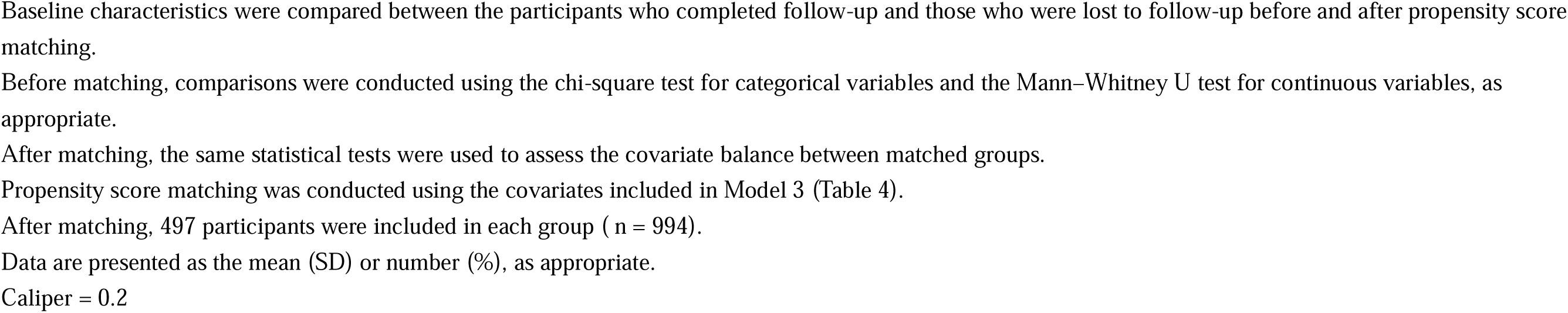
Baseline characteristics of participants who completed follow-up and those lost to follow-up, before and after propensity score matching.

**Supplementary Table 7.** Sex-stratified multivariate linear regression analysis of changes in MMSE scores after propensity score matching.

| Dependent variable | Sample | Model | $\beta$ for serum ATX (95% CI) | p-value |
| --- | --- | --- | --- | --- |
| ΔMMSE | all<br>(n=497) | model1 | 1.727 (0.407~3.046) | 0.0104* |
|  |  | model2 | 1.714 (0.347~3.081) | 0.0141* |
|  |  | model3 | 1.889 (0.509~3.269) | 0.0074* |
|  | male<br>(n=190) | model1 | 0.697 (-2.234~3.627) | 0.6396 |
|  |  | model2 | 0.052 (-3.025~3.130) | 0.9734 |
|  |  | model3 | 0.129 (-3.117~3.375) | 0.9374 |
|  | female<br>(n=307) | model1 | 2.032 (0.574~3.490) | 0.0065* |
|  |  | model2 | 2.416 (0.907~3.924) | 0.0018* |
|  |  | model3 | 2.728 (1.205~4.251) | 0.0005* |

Multivariate linear regression analyses were conducted using a propensity score-matched sample to examine the association between baseline serum ATX levels and longitudinal changes in the MMSE scores.
$\beta$ represents the unstandardized regression coefficient for serum ATX level.
Model 1 was adjusted for baseline age (in 2016), sex, serum ATX levels (in 2016), and baseline MMSE scores (in 2016).
Model 2 was further adjusted for ATX-related variables (liver fibrosis-FIB-3 Index, DHEAS score, obesity, and corticosteroid use).
Model 3 was further adjusted for dementia-related variables (education level > nine years, depression, hypertension, diabetes, history of stroke, living alone, drinking, smoking, and physical inactivity).
ΔMMSE was calculated as the MMSE score in 2022 minus the baseline MMSE score in 2016.

## Notes

### Author Declarations

Ethical approval for the baseline and follow-up surveys was obtained from the Ethics Committee of Kumamoto University, Kumamoto, Japan (Approval No. GENOME-333). Written informed consent was obtained from all participants or their legally authorized representatives. The present study analyzed stored biological samples and associated data as secondary research, which was separately approved by the same Ethics Committee (Approval No. RINRI-2915).

## References

1. Linnemann C, Lang UE. Pathways Connecting Late-Life Depression and Dementia. Front Pharmacol 2020; 11: 279.

2. Untu I, Davidson M, Stanciu GD et al. Neurobiological and therapeutic landmarks of depression associated with Alzheimer’s disease dementia. Front Aging Neurosci 2025; 17: 1584607.

3. Ramesh S, Govindarajulu M, Suppiramaniam V, Moore T, Dhanasekaran M. AutotaxinlJLysophosphatidic Acid Signaling in Alzheimer’s Disease. Int J Mol Sci 2018; 19.

4. Aoki J, Inoue A, Okudaira S. Two pathways for lysophosphatidic acid production. Biochim Biophys Acta 2008; 1781: 513–8.

5. Zhang X, Li M, Yin N, Zhang J. The Expression Regulation and Biological Function of Autotaxin. Cells 2021; 10.

6. Jose A, Kienesberger PC. Autotaxin-LPA-LPP3 Axis in Energy Metabolism and Metabolic Disease. Int J Mol Sci 2021; 22.

7. Castilla-Ortega E, Sánchez-López J, Hoyo-Becerra C et al. Exploratory, anxiety and spatial memory impairments are dissociated in mice lacking the LPA1 receptor. Neurobiol Learn Mem 2010; 94: 73–82.

8. Moreno-Fernández RD, Pérez-Martín M, Castilla-Ortega E et al. maLPA1-null mice as an endophenotype of anxious depression. Transl Psychiatry 2017; 7: e1077.

9. Itagaki K, Takebayashi M, Abe H et al. Reduced Serum and Cerebrospinal Fluid Levels of Autotaxin in Major Depressive Disorder. Int J Neuropsychopharmacol 2019; 22: 261–269.

10. Omori W, Kano K, Hattori K et al. Reduced Cerebrospinal Fluid Levels of Lysophosphatidic Acid Docosahexaenoic Acid in Patients With Major Depressive Disorder and Schizophrenia. Int J Neuropsychopharmacol 2021; 24: 948–955.

11. Ikeda H, Yatomi Y. Autotaxin in liver fibrosis. Clin Chim Acta 2012; 413: 1817–21.

12. Rancoule C, Dusaulcy R, Tréguer K, Grès S, Attané C, Saulnier-Blache JS. Involvement of autotaxin/lysophosphatidic acid signaling in obesity and impaired glucose homeostasis. Biochimie 2014; 96: 140–3.

13. Reeves VL, Trybula JS, Wills RC et al. Serum Autotaxin/ENPP2 correlates with insulin resistance in older humans with obesity. Obesity (Silver Spring) 2015; 23: 2371–6.

14. Sumida H, Nakamura K, Yanagida K et al. Decrease in circulating autotaxin by oral administration of prednisolone. Clin Chim Acta 2013; 415: 74–80.

15. Meng G, Tang X, Yang Z et al. Dexamethasone decreases the autotaxin-lysophosphatidate-inflammatory axis in adipose tissue: implications for the metabolic syndrome and breast cancer. Faseb j 2019; 33: 1899–1910.

16. Maninger N, Wolkowitz OM, Reus VI, Epel ES, Mellon SH. Neurobiological and neuropsychiatric effects of dehydroepiandrosterone (DHEA) and DHEA sulfate (DHEAS). Front Neuroendocrinol 2009; 30: 65–91.

17. Takeshita RSC, Nguyen AT, Auger AP, Chung WCJ. Cortisol, DHEAS, and the cortisol/DHEAS ratio as predictors of epigenetic age acceleration. Biogerontology 2025; 26: 164.

18. Ninomiya T, Nakaji S, Maeda T et al. Study design and baseline characteristics of a population-based prospective cohort study of dementia in Japan: the Japan Prospective Studies Collaboration for Aging and Dementia (JPSC-AD). Environ Health Prev Med 2020; 25: 64.

19. Yesavage JA, Sheikh JI. Geriatric Depression Scale (GDS): Recent Evidence and Development of a Shorter Version. Clinical Gerontologist 1986; 5: 165–173.

20. Folstein MF, Folstein SE, McHugh PR. “Mini-mental state”. A practical method for grading the cognitive state of patients for the clinician. J Psychiatr Res 1975; 12: 189–98.

21. Association AP. *Diagnostic and Statistical Manual of Mental Disorders (DSM-IV)*. American Psychiatric Association, Washington, DC, 1994.

22. Association AP. Diagnostic and Statistical Manual of Mental Disorders, Third Revised Edition (DSM-III-R). American Psychiatric Association, Washington, DC, 1987.

23. Kariyama K, Kawanaka M, Nouso K et al. Fibrosis-3 Index: A New Score to Predict Liver Fibrosis in Patients With Nonalcoholic Fatty Liver Disease Without Age as a Factor. Gastro Hep Adv 2022; 1: 1108–1113.

24. Hidaka Y, Hashimoto M, Suehiro T et al. Association between choroid plexus volume and cognitive function in community-dwelling older adults without dementia: a population-based cross-sectional analysis. Fluids Barriers CNS 2024; 21: 101.

25. Desikan RS, Ségonne F, Fischl B et al. An automated labeling system for subdividing the human cerebral cortex on MRI scans into gyral based regions of interest. Neuroimage 2006; 31: 968–80.

26. Adam EK, Kumari M. Assessing salivary cortisol in large-scale, epidemiological research. Psychoneuroendocrinology 2009; 34: 1423–36.

27. Méaux MN, Regnier M, Portefaix A et al. Circulating autotaxin levels in healthy teenagers: Data from the Vitados cohort. Front Pediatr 2023; 11: 1094705.

28. Nakamura K, Igarashi K, Ide K et al. Validation of an autotaxin enzyme immunoassay in human serum samples and its application to hypoalbuminemia differentiation. Clin Chim Acta 2008; 388: 51–8.

29. Yung YC, Stoddard NC, Mirendil H, Chun J. Lysophosphatidic Acid signaling in the nervous system. Neuron 2015; 85: 669–82.

30. Herr DR, Ong JH, Ong WY. Potential Therapeutic Applications for Inhibitors of Autotaxin, a Bioactive Lipid-Producing Lysophospholipase D, in Disorders Affecting the Nervous System. ACS Chem Neurosci 2018; 9: 398–400.

31. Debruyne H, Van Buggenhout M, Le Bastard N et al. Is the geriatric depression scale a reliable screening tool for depressive symptoms in elderly patients with cognitive impairment? Int J Geriatr Psychiatry 2009; 24: 556–62.

32. Tanaka M, Okudaira S, Kishi Y et al. Autotaxin stabilizes blood vessels and is required for embryonic vasculature by producing lysophosphatidic acid. J Biol Chem 2006; 281: 25822–30.

33. Hahnefeld L, Hackel J, Trautmann S et al. Healthy plasma lipidomic signatures depend on sex, age, body mass index, and contraceptives but not perceived stress. Am J Physiol Cell Physiol 2024; 327: C1462–c1480.

34. Moriyama R, Fukushima N. Expression of lysophosphatidic acid receptor 1 in the adult female mouse pituitary gland. Neurosci Lett 2021; 741: 135506.

35. Sánchez-Marín L, Jiménez-Castilla V, Flores-López M et al. Sex-specific alterations in emotional behavior and neurotransmitter systems in LPA(1) receptor-deficient mice. Neuropharmacology 2025; 268: 110325.

36. Frugier T, Crombie D, Conquest A et al. Modulation of LPA receptor expression in the human brain following neurotrauma. Cell Mol Neurobiol 2011; 31: 569–77.

37. Aston C, Jiang L, Sokolov BP. Transcriptional profiling reveals evidence for signaling and oligodendroglial abnormalities in the temporal cortex from patients with major depressive disorder. Mol Psychiatry 2005; 10: 309–22.

38. Nagy C, Maitra M, Tanti A et al. Single-nucleus transcriptomics of the prefrontal cortex in major depressive disorder implicates oligodendrocyte precursor cells and excitatory neurons. Nat Neurosci 2020; 23: 771–781.

